# Association between pharmacological tenofovir adherence measures and subsequent 24-week viral load outcomes for people living with HIV in South Africa

**DOI:** 10.1101/2025.11.10.25339878

**Authors:** Nicola Bodley, Katya Govender, Pravikrishnen Moodley, Natasha Samsunder, Yukteshwar Sookrajh, Philip J Turner, Christopher C Butler, Gail N Hayward, Monica Gandhi, Paul K. Drain, Nigel Garrett, Jienchi Dorward

## Abstract

**Background:** Objective measures of antiretroviral therapy (ART) adherence, such as tenofovir (TFV) concentrations, may allow for targeted interventions to prevent future HIV viraemia. We evaluated three TFV adherence measures and their association with current and 24-week viral load (VL) outcomes.

**Methods:** In a sub-study of the South African-based POwER trial, we measured urine TFV using a point-of-care (POC) antibody-based assay and two metrics assessed via liquid-chromatography-mass-spectrometry: quantitative urine TFV, and TFV-diphosphate (TFV-DP) concentrations in dried blood spots (DBS). We assessed the association between current and subsequent 24-week VL outcomes and these three measures using logistic regression models. We also compared the baseline characteristics of individuals with detectable vs. undetectable POC urine TFV results at enrolment.

**Results:** Of 124 participants, 54.8% female, median age 39 years, 101 (81.5%) had detectable POC urine TFV and 23 (18.5%) did not. Higher TFV-DP concentrations in DBS were negatively associated with viraemia after 24-weeks (OR 0.83, 95% CI 0.725-0.928, p=0.003), whilst a detectable POC urine TFV (OR 0.62, 95% CI 0.22-1.82, p=0.380) and quantitative urine TFV (OR 0.98, 95% CI 0.95-1.00, p=0.153) showed no significant association. Compared to those with detectable POC urine TFV, those with undetectable POC urine TFV were more likely to be concurrently viraemic at enrolment (78.3% vs 25.7%, p<0.001) and have a CD4 count <200 cells/uL (34.8% vs 12.9%, p=0.001).

**Conclusion:** DBS TFV-DP was associated with viraemia at 24-weeks and could help predict future viraemia. Undetectable POC urine TFV was associated with recent ART initiation, current viraemia, and lower CD4 count.

**Trial registration:** Pan African Clinical Trials Registry (PACTR202001785886049) and the South African Clinical Trials Registry (DOH-27-072020-6890).

## Introduction

Lifelong adherence to antiretroviral therapy (ART) is crucial for people living with HIV (PLWH), but assessing and supporting adherence is challenging. Viral load (VL) measurement is the primary marker of treatment outcomes, and in South Africa (SA) an undetectable VL <50 copies/mL is defined as treatment success (1). However, VL monitoring is costly, cannot differentiate between poor adherence and viral resistance, and relies on centralized laboratories, with delayed results hindering timely interventions. Furthermore, measuring VL allows detection of established viraemia, leaving no time to intervene before viraemia occurs. Measuring ART drug concentrations can objectively assess adherence and identify PLWH who require early intervention before viraemia arises.

In SA, current (2023) HIV treatment guidelines recommending a fixed combination of tenofovir disoproxil fumarate (TDF), lamivudine and dolutegravir (TLD) for first-line ART (2). Dolutegravir is well tolerated, effective in achieving and maintaining viral suppression, and has a high genetic barrier to resistance (3–4). The fixed dose combination of TLD means that measures of tenofovir (TFV) concentrations may be particularly useful for measuring overall adherence to this widely used single tablet combination.

Both short- and long-term measures of TFV adherence exist. Tenofovir-diphosphate (TFV-DP) concentrations measured in dried blood spots (DBS) by liquid chromatography-tandem mass spectrometry (LC-MS/MS) are associated with TDF exposure in the preceding six to eight weeks (5), and can predict future viraemia, however this technology still requires samples to be sent to and processed in laboratories (5–8). More clinically accessible methods of TFV assessment are needed, such as antibody-based point-of-care (POC) urine TFV assays which provide a measure of recent TDF exposure (preceding three to four days), giving insight into recent or short-term adherence (9). POC-TFV assays can provide results within minutes in a clinic setting and are straightforward to use with minimal training (10). Use of POC TFV assays in combination with enhanced adherence counselling (EAC) has already been associated with high levels of resuppression after virological failure (11). Furthermore, POC-TFV, quantitative urine TFV by LC-MS/MS and TFV-DP are associated with concurrent viraemia (12–13). However, whether POC-TFV and quantitative urine TFV measures are associated with future viraemia has not been demonstrated.

This study aimed to determine whether DBS TFV-DP concentrations, quantitative urine TFV concentrations, and POC-TFV, are associated with current and subsequent 24-week VL values. It also aimed to compare the baseline characteristics of individuals in whom the urine POC-TFV assay was detectable versus undetectable.

## Methods

### Study design

We conducted a sub-study within the POwER trial, a feasibility study comparing POC VL testing with standard laboratory VL testing among people with recent viraemia, which has been described previously (14,15).

### Setting

The study took place at an urban and a rural clinic in KwaZulu-Natal, SA, which are supported by the Centre for the AIDS Programme of Research in South Africa (CAPRISA).

### Participants

We included all POwER participants who were taking TDF. PLWH were eligible for POwER if they were receiving first-line dolutegravir or efavirenz-based ART and with recent viraemia >1000 copies/mL in the past 6 weeks, for which they had not yet received enhanced adherence counselling (EAC).

### Procedures

Consent for this sub-analysis was included in the original POwER consent. Baseline demographic, behavioural, and clinical characteristics of participants were collected at enrolment, EAC was conducted and participants had urine, whole blood and plasma samples taken and stored at −80°C, for retrospective testing. Participants were randomized to receive either POC or standard laboratory-based repeat VL testing after 12 weeks, with the primary outcome VL measured 24-weeks after enrolment. The VL measured at enrolment is considered the concurrent VL in this study.

### Laboratory testing

Urine was thawed and sent for quantitative urine TFV adherence testing using LC-MS/MS at the Africa Health Research Institute (AHRI) in Durban, and for POC-TFV assay (Abbott Laboratories, Abbott Park, IL, USA) testing at the CAPRISA Clinical Trial Unit Laboratory.

The Abbot POC-TFV assay is an antibody-based test for TFV that utilizes Lateral Flow Immunoassay (LFA) technology and was previously validated to detect a TFV concentration of ≥1500 ng/ml against the reference standard LC-MS/MS (16). This threshold determines whether a person has taken at least one dose of TDF within the prior four days. Specifically, 3-4 urine drops were added to the test well, with the result read by two independent laboratory technicians after 3-5 minutes. Photos of discrepant results were adjudicated by a third investigator. For TFV-DP measurement, DBS were prepared from EDTA specimens and stored at −80°C before being thawed and analysed using LC-MS/MS at AHRI (supplement). 24-week VL was measured in thawed plasma samples on the cobas 6800 platform (Roche, Basel, Switzerland) in the National Health Laboratory Service at the Inkosi Albert Luthuli Hospital in Durban.

### Exposures and outcomes

For the primary analysis, the exposures of interest were POC-TFV, DBS TFV-DP, and quantitative urine TFV, with the primary outcome being viraemia (VL>50 copies/mL) at 24-weeks.

For the secondary analysis, the exposures of interest were baseline sociodemographic and laboratory (including current VL and CD4 count) factors, and the outcome of interest was detectable vs. undetectable urine POC-TFV.

### Statistical analysis

For the primary analysis, we used separate univariable logistic regression models to analyze the relationship between each of the measures of adherence (binary POC-TFV [detected versus not-detected], quantitative urine TFV and quantitative DBS TFV-DP as independent exposures) and the 24-week viraemia outcome. In a sensitivity analysis, we included an interaction term between the TFV measures and a binary exposure variable of baseline ART regimen (dolutegravir or efavirenz-based), to evaluate whether any associations differed by ART regimen.

In a secondary analysis, we described baseline clinical and laboratory characteristics of participants with detectable versus undetectable TFV in the urine, using the POC TFV assay, and compared the proportions using chi-squared tests.

### Ethical approval

POwER was approved by the University of KwaZulu-Natal Biomedical Research Ethics Committee (BREC00000836/2019) and the University of Oxford Tropical Research Ethics Committee (OxTREC66-19).

## Results

### Study Population

124 participants taking TDF were enrolled into POwER between 20 August 2020 and 25 March 2022. Median age was 39.0 years (interquartile range (IQR) 34.0 to 45.0 years, Table 1) and 68/124 (54.8%) were women. Participants had been on ART a median of 4.1 years (IQR 1.5 to 6.1) and had been on their current regimen (40.3% dolutegravir, 59.7% efavirenz-based) for a median 1.6 years (IQR 0.7 to 5.0). Participants presented a median 15 days (IQR 12.8 to 21.0) since their pre-enrolment viraemic result, that determined their eligibility for POwER.

**Table 1:** Baseline characteristics of participants stratified by urine TFV results.

| Characteristic | Variable | Total | TFV Not Present (n=23) | TFV Present (n=101) | p-value |
| --- | --- | --- | --- | --- | --- |
| Age, years | Median (IQR) | 39.0 (34.0 to 45.0) | 38.0 (34.0 to 51.0) | 39.0 (34.0 to 45.0) | 0.938 |
| Gender | Female | 68 (54.8) | 9 (39.1) | 59 (58.4) | 0.108 |
|  | Male | 56 (45.2) | 14 (60.9) | 42 (41.6) |  |
| Time since ART initiation, years | Median (IQR) | 4.1 (1.5 to 6.1) | 2.0 (0.7 to 4.4) | 4.3 (2.1 to 6.2) | 0.002 |
| Time on current regimen, years | Median (IQR) | 1.6 (0.7 to 5.0) | 0.8 (0.5 to 2.1) | 2.1 (0.8 to 5.2) | 0.017 |
| Time on dolutegravir-based regimen, years | Median (IQR) | 0.6 (0.5 to 1.0) | 0.6 (0.5 to 1.1) | 0.6 (0.5 to 1.0) | 0.807 |
| Current TB | No | 122 (98.4) | 23 (100.0) | 99 (98.0) | 1.000 |
|  | Yes | 2 (1.6) |  | 2 (2.0) |  |
| ART side effects | No | 120 (96.8) | 23 (100.0) | 97 (96.0) | 1.000 |
|  | Yes | 4 (3.2) |  | 4 (4.0) |  |
| Last time participant missed a dose of ART | <2 weeks | 30 (24.2) | 11 (47.8) | 19 (18.8) | 0.014 |
|  | 2-4 weeks | 16 (12.9) | 3 (13.0) | 13 (12.9) |  |
|  | 1-3 months | 16 (12.9) | 4 (17.4) | 12 (11.9) |  |
|  | >3 months | 8 (6.5) | 1 (4.3) | 7 (6.9) |  |
|  | Never | 54 (43.5) | 4 (17.4) | 50 (49.5) |  |
| Number of ART doses missed in past 4 days | 0 | 95 (76.6) | 11 (47.8) | 84 (83.2) | <0.001 |
|  | 1 | 14 (11.3) | 3 (13.0) | 11 (10.9) |  |
|  | 2 | 9 (7.3) | 5 (21.7) | 4 (4.0) |  |
|  | 3 | 2 (1.6) | 1 (4.3) | 1 (1.0) |  |
|  | 4 | 4 (3.2) | 3 (13.0) | 1 (1.0) |  |
| Ethnicity | Black African | 121 (97.6) | 22 (95.7) | 99 (98.0) | 0.463 |
|  | Coloured | 1 (0.8) |  | 1 (1.0) |  |
|  | Other | 2 (1.6) | 1 (4.3) | 1 (1.0) |  |
| Enrolment CD4 count category, cells/μL | <200 | 21 (16.9) | 8 (34.8) | 13 (12.9) | 0.001 |
|  | 200-349 | 25 (20.2) | 7 (30.4) | 18 (17.8) |  |
|  | 350-499 | 29 (23.4) | 6 (26.1) | 23 (22.8) |  |
| | ≥500 | 49 (39.5) | 2 (8.7) | 47 (46.5) | |
| Time since pre-enrolment viral load, days | Median (IQR) | 15.0 (12.8 to 21.0) | 15.0 (13.5 to 18.0) | 14.0 (12.0 to 21.0) | 0.747 |
| ART regimen at enrollment | TDF / 3TC / DTG | 49 (39.5) | 12 (52.2) | 37 (36.6) | 0.237 |
|  | TDF / FTC / - | 75 (60.5) | 11 (47.8) | 64 (63.4) |  |
| Urine tenofovir concentration, ng/mL | Median (IQR) | 20000 (7280 to 33625) | 207 (0.0 to 987) | 23100 (14400 to 37100) | <0.001 |
| DBS tenofovir diphosphate concentration, fmol/sample | Median (IQR) | 734.2 (471.0 to 1015.2) | 263.4 (40.2 to 429.9) | 814.7 (576.4 to 1162.5) | <0.001 |
| Enrolment viral load | <50 copies/mL | 57 (46.0) | 2 (8.7) | 55 (54.5) | <0.001 |
|  | 50-999 copies/mL | 23 (18.5) | 3 (13.0) | 20 (19.8) |  |
|  | ≥1000 copies/mL | 44 (35.5) | 18 (78.3) | 26 (25.7) |  |

During the initial phase of enrolment into the POwER study, we were informed that 45 participants had their pre-enrolment VLs measured on a defective VL analyzer which overestimated some VL results meaning that part of the “viraemic” cohort enrolled into the study were virally suppressed. After being informed, all the affected participants were agreeable to continue in POwER and were included in this sub-study in order to have participants with a history of both viraemia and viral suppression.

Of the 124 participants, 100 (81.5%) had a detectable urine POC-TFV result, and 23 (18.5%) an undetectable result (**Table 1**). Overall, the median quantitative urine TFV concentration was 20000 ng/mL (IQR 7280 to 33625) and median TFV-DP concentration was 734.2 fmol/sample (IQR 471.0 to 1015.2).

### Association between TFV and TFV-DP measures and subsequent 24-week VL outcomes

One participant did not have a 24-week VL, despite attending the 24-week visit, and was excluded from the primary analysis. Four (3.2%) were lost to follow-up (missed week 24 visit) and were assumed to have an unsuppressed 24-week VL.

We did not find evidence that the POC-TFV assay (odds ratio [OR] 0.622, 95% CI 0.222-1.920, p=0.380, Table 2) or quantitative urine TFV concentrations (OR 0.982, 95% CI 0.954-1.000, p=0.153) were associated with 24-week viraemia. In contrast, DBS TFV-DP concentrations were strongly associated with 24-week viraemia (OR 0.828, 95% CI 0.725-0.928, p=0.003), meaning for every 100 fmol/punch increase in TFV-DP the odds of viraemia decreased by 0.83.

**Table 2:** Logistic regression models of the association between point-of-care urine tenofovir results, quantitative urine tenofovir concentrations, and dried blood spot tenofovir diphosphate concentrations, and subsequent 24-month viraemia.

|  | Overall |  | Dolutegravir-based ART |  | Efavirenz-based ART |  |
| --- | --- | --- | --- | --- | --- | --- |
| Logistic regression models |  |  |  |  |  |  |
|  | Odds ratio<br>(95% CI) | P | Odds ratio<br>(95% CI) | P | Odds ratio<br>(95% CI) | P |
| Viraemia ≥50 copies/mL |  |  |  |  |  |  |
| Positive point-of-care urine TFV test | 0.62<br>(0.22-1.92) | 0.380 | 0.96<br>(0.23-5.04)* | 0.962 | 0.44<br>(0.10-2.31)* | 0.290 |
| Urine TFV concentration (increase of 1000 ng/mL) | 0.98<br>(0.95-1.00) | 0.153 | 0.97<br>(0.93-1.00)<sup>†</sup> | 0.121 | 0.99<br>(0.96-10.3)<sup>†</sup> | 0.749 |
| DBS TFV-DP concentration (increase of 100 fmol/punch) | 0.83<br>(0.73-0.93) | 0.003 | 0.840<br>(0.68-1.00)<sup>‡</sup> | 0.079 | 0.82<br>(0.68-0.96)<sup>‡</sup> | 0.024 |

\* LRT for interaction p = 0.477, <sup>†</sup>LRT for interaction p = 0.362, <sup>‡</sup>LRT for interaction p = 0.854

### The effect of baseline ART regimen on the association between TFV and TFV-DP measures and subsequent 24-week VL outcomes

In the sensitivity analysis testing for effect modification by baseline ART regimen (dolutegravir-versus efavirenz-based ART) (**Table 2)** there was no evidence that the baseline ART regimen modified the association between the subsequent 24-week VL outcomes and POC-TFV (likelihood ratio test for interaction [LRT] p=0.477), quantitative urine TFV (LRT p=0.362) or DBS TFV-DP (LRT p=0.854).

### Characteristics of study participants stratified by POC urine TFV results

There was no significant difference between people with detectable and undetectable POC-TFV results based on age, sex, ethnicity, ART regimens or the presence of reported ART side effects. Compared to participants with detectable TFV, those with undetectable TFV, had been on ART a median of 2.0 (IQR 0.7-4.4) years versus 4.3 (IQR 2.1-6.2) years (p=0.002) and on their current regimen a median of 0.8 (IQR 0.5-2.1) years versus 2.1 (IQR 0.7-5.0) years (p=0.017). At enrolment, a greater proportion of participants with undetectable TFV had a CD4 count <200 cells/uL compared to those with detectable TFV (34.8% vs 12.9% p=0.001) and a higher proportion had concurrent viraemia (VL ≥1000 copies/mL) (78.3% vs 25.7% p<0.001).

## Discussion

We found that DBS TFV-DP concentrations were associated with subsequent 24-week viraemia. While the odds ratios for POC-TFV and quantitative urine TFV for future viraemia were less than one, our sample size was small and the confidence intervals were broad and included one. We also found that those with undetectable TFV were more likely to be earlier in their ART journey, have lower CD4 counts, and have current viraemia.

DBS TFV-DP has been shown to correlate with HIV treatment outcomes but requires centralized laboratory processing (5-8,17-18,), while POC-TFV assays may be used in clinics. Although other studies have shown the relationship between POC-TFV and concurrent viremia, ours is the only study, to our knowledge, to evaluate the relationship between POC-TFV results and future VL outcomes.

In our study, we found no association between POC-TFV and quantitative urine TFV results and subsequent 24-week viraemia. This may be due to our relatively small sample size, or because these are measures of short-term adherence with a weaker relationship with future viraemia than TFV-DP in DBS which measures longer term adherence. We previously showed that POC-TFV results were associated with concurrent viraemia measured at the same timepoint (12). Therefore, the POC-TFV assay, which has been shown to be acceptable in pre-exposure prophylaxis studies (19) and recent qualitative studies in SA (10, 20), may be useful in public primary healthcare clinics for rapidly detecting poor adherence and guiding counselling for PLWH. Adherence interventions with POC-TFV guided feedback have been associated with VL improvements and are being more widely studied (11). Implementation studies and clinical trials are evaluating POC-TFV assays among PLWH in SA, and the updated SA ART guidelines allow adherence assessment using ART measurement in blood or urine, which could assist with targeting HIV drug resistance testing or triaging of PLWH into differentiated models of care (2).

Our finding that people with a negative POC-TFV assay result were more likely to be at an earlier stage of ART, have lower CD4 cell counts, and are more likely to have current viraemia suggesting that early timepoints are critical for PLWH, and more focus should be placed on providing early adherence support after ART initiation. In addition to baseline CD4 count in other studies age and sex are important predictors of future treatment failure (21).

This study’s strengths lie in its comparative analysis of three different pharmacological measures of TFV adherence and its focus on future viraemia as opposed to current viraemia. The study took place in SA, a high disease burden setting, which would benefit from timely cost-effective strategies to assess and manage adherence.

Limitations include a small sample size, with data collected from only two sites, and the relatively short follow-up period, meaning larger, longer-term studies are required.

Measuring TFV-DP in DBS is strongly associated with subsequent 24-week viraemia, in both people receiving dolutegravir- or efavirenz-based ART and could provide an opportunity to target adherence interventions, reduce viraemia and prevent drug resistance. The association of undetectable POC-TFV with current viraemia and lower CD4 counts earlier after ART initiation suggests future interventions to avert poor outcomes.

## Data Availability

All data produced in the present study are available upon reasonable request to the authors

## Author contributions

JD, NB and NG conceived the study. KG, PM, and NS were responsible for laboratory testing. YS, PT, CB and GH contributed to study design. JD analysed the data. NB wrote the first draft of the manuscript. All authors critically reviewed and edited the manuscript and consented to final publication.

